# Age-Related Macular Degeneration, Cardiovascular Disease and Stroke

**DOI:** 10.1101/2021.09.13.21263389

**Authors:** Gerardo Ledesma-Gil, Oscar Otero-Marquez, Sharmina Alauddin, Yuehong Tong, Wei Wei, Katy Tai, Harriet Lloyd, Micaela Koci, Catherine Ye, Cinthi Pillai, Maria Scolaro, Arun Govindaiah, Alauddin Bhuiyan, Avnish Deobhakta, Richard B. Rosen, Lawrence A. Yannuzzi, K. Bailey Freund, R. Theodore Smith

**Affiliations:** Vitreous Retina Macula Consultants of New York, New York, USA; Department of Ophthalmology, New York Eye and Ear Infirmary of Mount Sinai, New York, New York, USA; Retina Department, Institute of Ophthalmology Fundación Conde de Valenciana, Mexico City, Mexico; Icahn School of Medicine at Mount Sinai, New York, New York, USA; Department of Neurology, NYU Grossman School of Medicine. New York, New York, USA; Rutgers New Jersey Medical School, Newark, New Jersey, USA; iHealthscreen Inc., New York, USA

**Keywords:** Age related macular degeneration, reticular macular disease, stroke, cardiovascular disease, risk factors, subretinal drusenoid deposits, drusen, choroid

## Abstract

**Importance:** High-risk vascular diseases (HRVs) may remain undetected until catastrophe ensues. Detection from non-invasive retinal imaging would be highly significant.

**Objective:** To demonstrate that certain lesions of Age-Related Macular Degeneration (AMD) found on retinal imaging correlate with co-existing HRVs.

**Design:** Cross-sectional cohort study. Two years. Retinal image graders blinded to HRV status.

**Setting:** 2 retina referral clinics.

**Participants:** 151 consecutive AMD patients, ages 50-90, 97 females, 54 males, with lesions of soft drusen and/or subretinal drusenoid deposits (SDD). 12 others approached, 10 refused, 2 excluded.

**Methods:** Patients were classified by retinal imaging into SDD (SDD present, +/- drusen) or nonSDD (soft drusen only), and by history into HRV (cardiac pump defect (myocardial infarction (MI), coronary artery bypass grafting (CABG), congestive heart failure (CHF)), valve defect, and carotid stroke) or nonHRV, with serum risk factors and medical histories.

**Main Outcome Measures:** Correlations of HRV with SDD and other covariates (Univariate chi-square and multivariate regression). Performance of Machine Learning predicting HRV.

**Results:** 75 SDD subjects; 76 nonSDD subjects; HRV prevalence 19.2% (29/151).

1. High density lipoprotein (HDL) < 62 mg/Dl was found in 24/29 HRV, 42/122 nonHRV, OR 12.40, 95% Confidence Interval (CI) 5.125-30.014; p= 0.0002.
2. 15 Pump defects, 14/15 SDD, 8 Valve defects, 6/8 SDD (4 severe aortic stenosis), 6 carotid strokes, 5/6 SDD. Total HRVs 29, 25/29 SDD, OR 9.0, 95% CI 2.95-27.46; p= 0.000012.
3. Adjusted multivariate correlations. HRV with SDD (p= 0.000333). SDD and HDL < 62 with HRV (p= 0.000098 and 0.021).
4. Machine Learning prediction of HRVs from SDD status and HDL level: specificity 87.4%, sensitivity 77.4%, accuracy 84.9%; 95% CIs(%) 79.0-93.3, 58.0-90.4, 77.5-90.7, respectively.

**Conclusions and Relevance:** High-risk vascular diseases were accurately identified in a cohort of AMD patients from the presence of characteristic deposits (SDDs) on imaging and HDL levels. The SDDs are directly consequent to inadequate ocular perfusion resulting from the systemic vasculopathies. Further validation in larger cohorts of both vasculopathic and AMD subjects could bring this system into widespread medical practice, to reduce mortality and morbidity from vascular disease, particularly in women, where undiagnosed cardiac disease remains a serious issue.

**Key Points:** *Question:* **What is the relationship and driving mechanism between** High Risk Vascular **Diseases (HRVs) and Age-Related Macular Degeneration (AMD)?**

*Findings:* The specific AMD lesions of Subretinal Drusenoid Deposits (SDDs) were found to be highly correlated with and directly consequent to the inadequate ocular perfusion resulting from the HRVs of severe cardiac pump insufficiency or valve defect, and carotid occlusion, These vasculopathies could be predicted from the presence of SDDs on spectral domain optical coherence tomography (SD-OCT) imaging and serum HDL.

*Meaning:* Screening for SDDs with SD-OCT imaging could reduce mortality and morbidity from severe vascular disease, particularly in women, where undiagnosed cardiac disease remains a serious issue.

---

Cardiovascular disease (CVD) and stroke are the leading causes of death in the developed world.^1^ Age related macular degeneration (AMD) is the leading cause of blindness,^2^ However, despite lipid-rich lesions in both and some common risk factors, decades of study have failed to find consistent associations, e.g., Stroke, no association; myocardial infarction (MI), positive and Coronary Artery Disease (CAD), inverse associations,^3-5^ and in metanalyses considering AMD and CVD (Wang et al, N = 29,964,334) and AMD and stroke (Fernandez et al, N = 1,420,978), no associations .^6,7^ We present herein a strong association between a specific lesion of AMD and a specific subset of high-risk vascular diseases (HRVs) driven by a clear mechanism that not only resolves this long-standing puzzle, but also can predict these diseases from retinal imaging.

The two lipid-laden lesions of AMD are soft drusen, beneath, and subretinal drusenoid deposits (SDD) above, the retinal pigmented epithelium (RPE),^8^ the nourishing layer beneath the outer retina where AMD develops, in turn supplied by the choroidal vasculature/choriocapillaris (CC).^9^ Both deposits can progress to advanced AMD,^2,10,11^ and SDD, aka reticular pseudodrusen (RPD),^12^ confer twice the risk.^13,14,15^ CC insufficiency uniformly accompanies SDD on angiography^16^ and non-invasive optical coherence tomography angiography (OCTA), often with choroidal thinning, additionally suggesting loss of choroidal volume.^17^

Together, CC insufficiency and SDDs comprise the AMD phenotype of Reticular Macular Disease (RMD).^16^

## Hypotheses

I. RMD and its SDDs are linked strongly to high risk vascular disorders (HRVs) of cardiac pump deficiency and valve defect, and carotid stroke. II. The mechanism is choriocapillaris (CC) insufficiency driven by the systemic vasculopathy. III, RMD can predict co-existing HRV. This new paradigm, built precisely on RMD,^13-15^ should facilitate timely diagnosis and intervention in HRVs.

Identifying drusen and the SDD lesions of RMD depends critically on retinal imaging: soft drusen, easily with color fundus photography (CFP);^18^ SDD, more difficult, first accomplished with blue light.^19^ (**Figure 1A)**. Although Arnold et al then showed major SDD associations with advanced AMD,^20^ acceptance was discouraged.^21^

**Figure 1.**
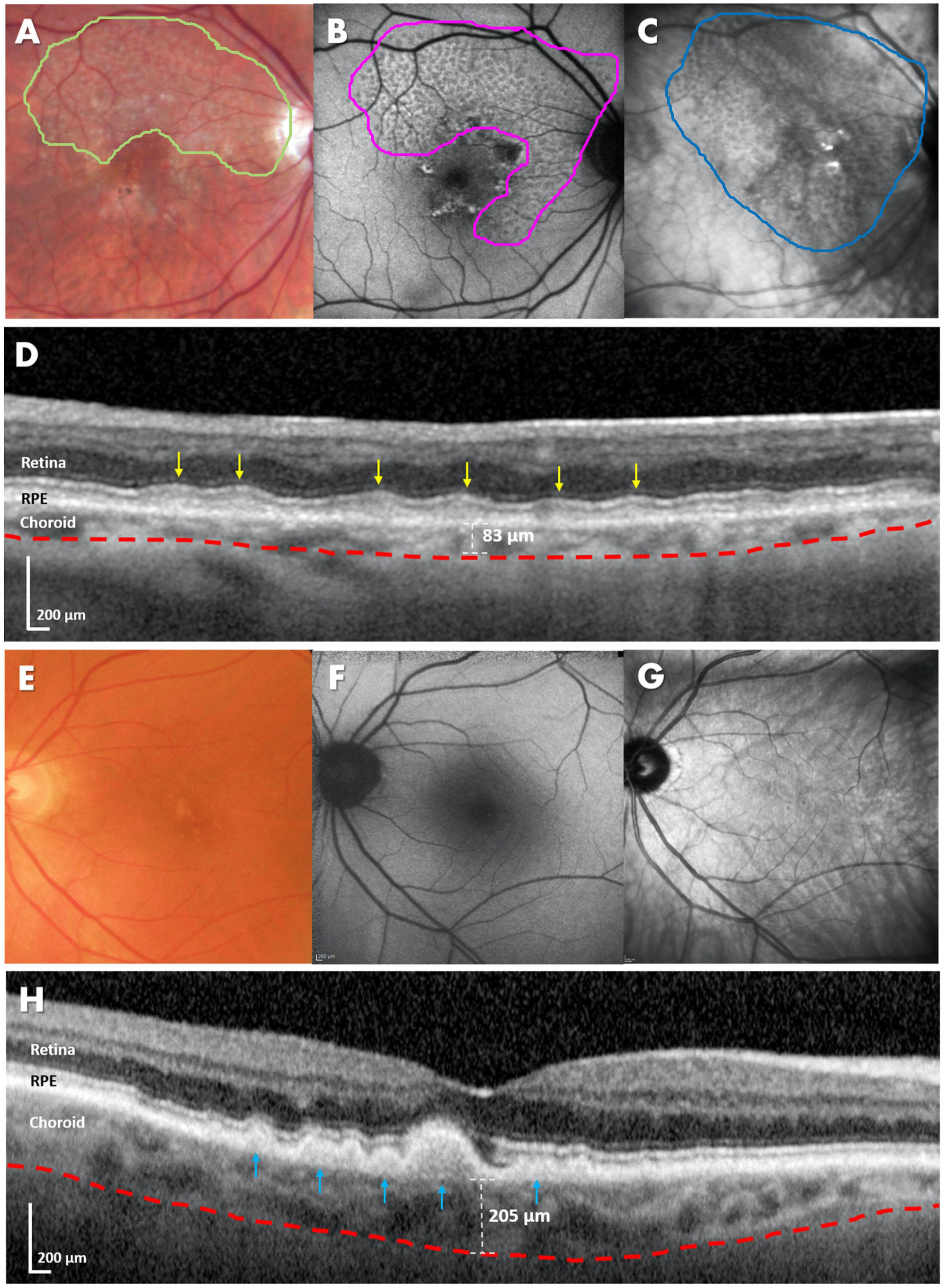
Multimodal Imaging and spectral domain optical coherence tomography (SD-OCT) imaging of Reticular Macular Disease (RMD) and Soft Drusen in AMD. **A-D) 81-85 year-old age range WF with RMD. A) Color fundus photography (CFP)** is inefficient for imaging RMD. This photo required contrast enhancement to visualize the characteristic reticular pale flecks of RMD (green border). **B) Autofluorescence (AF)** lesions of RMD are fairly homogeneous and moderately hypo-autofluorescent (magenta border). **C) Near infrared reflectance (NIR)** lesions of RMD are fairly homogeneous and moderately hypo-reflectant (blue border). The en face multi-modal images all show fairly homogeneous lesions in compact distributions with well-defined borders, most prevalent superiorly. Capture of RMD lesions is typically most efficient with AF/NIR and least with CFP, as here, with multiple AF and NIR lesions that are pathognomonic for RMD. **D) SD-OCT Imaging**. RMD on SD-OCT consists of subretinal drusenoid deposits (SDD) and CC insufficiency on SD-OCT angiography (SD-OCTA), usually accompanied by choroidal thinning. SDD are defined by the presence of hyperreflective material between the retinal pigment epithelium (RPE) and ellipsoid zone (EZ). Stage 2 lesions are large enough to distort the EZ on the SD-OCT. The EZ is the wavy white line above the flat line of the RPE. Between the EZ and the RPE are seen the mounds, also wave-like, of SDD causing the distortion (yellow arrowheads). Smaller Stage 1 lesions not distorting the EZ can be seen between the Stage 2 SDD. This is pure RMD (no soft drusen; see soft drusen, **Panel D)**. The choroidal-scleral interface follows the red dashed line. Choroidal thickness, measured from the base of the RPE to this interface is 83 microns, significantly thinner than for age-matched normals. **E-H) 76-80 year-old age range WF with Soft drusen. E) Color fundus photography (CFP)** shows yellowish confluent lesions close to the fovea. **F) Autofluorescence (AF)** lesions of soft drusen have a slightly hyperautofluorescent or neutral appearance. **G) Near infrared reflectance (NIR)** lesions of soft drusen have a hyperreflectant appearance. **H) SD-OCT Imaging**. Soft drusen are focal deposits of extracellular debris located between the basal lamina of the RPE and the inner collagenous layer of Bruch membrane (BrM), seen on SD-OCT as lumps of intermediate reflectivity on BrM elevating the bright line of the RPE (blue arrowheads). BrM is seen intermittently as a thin white line under the drusen where the RPE is elevated. BrM cannot be seen on standard OCT with RPE flat, **panel D**. Note the difference in choroidal thickness between the two phenotypes, with the choroid being thinner in RMD (83 microns vs 205 microns in the soft drusen phenotype).

Finally, scanning laser ophthalmoscopy (SLO), with autofluorescence (AF) and near-infrared reflectance (NIR) imaging, easily identified the SDD^16^ as hypoautofluorescent on AF^22^ and hyporeflectant on NIR^16^ (**Figure 1B, C)**. Histopathological identification of SDD^12^ in the subretinal space, with drusen in the sub-RPE space, confirmed *in vivo* by spectral domain OCT (SD-OCT),^23^ with choroidal thinning commonly found with SDD (**Figure 1D)** completed the structure of RMD. SDD/CC insufficiency and soft drusen (**Figure 1**) are the respective structural/functional markers for the distinct RMD and drusen pathways of AMD.

The choroidal vascular associations of SDD suggested our Hypotheses: the CC insufficiency of RMD is in large part the sequelae of systemic vascular disorders, whose structural biomarkers are SDD. Indeed, the association of coronary artery disease (CAD) with SDD/RMD was shown in a smaller case-control study,^10^ as was the association of CAD with generalized choroidal thinning,^11^ since confirmed, with OCTA documentation of CC insufficiency.^24^

In this paradigm, the common underlying mechanism is poor perfusion of the ophthalmic artery (OA). Indeed, diminished flow in the OA has recently been demonstrated by magnetic resonance angiography (MRA) in AMD generally versus controls, with significant further reduction in advanced AMD ty. Although this study did not differentiate between SDD and nonSDD subjects, and did not correlate flow measurements to vascular disease, the findings are quite consistent with our hypothesis.

The OA is too small for atherosclerotic disease (ASD) *per se*, the main lesion of CAD and stroke. However, perfusion of the OA, the first intracranial branch of the internal carotid artery (ICA), is vulnerable to plaque at the carotid bifurcation. Thus, carotid stroke is a high probability marker for unilateral OA compromise (**Figure 2**, left-sided stroke, ipsilateral ICA occlusion and RMD). Bilateral RMD can follow compromised cardiac output of any etiology. CAD is such a cause^10^ (**Figure 3**, bilateral pure RMD (no drusen) consequent to MI). However, there are other causes of decreased cardiac output such as congestive heart failure (CHF) from any cause and severe valvular disease. Valve disease, to our knowledge, has not been previously reported in association with AMD (**Figure 4, pure** RMD consequent to severe aortic stenosis).

**Figure 2.**
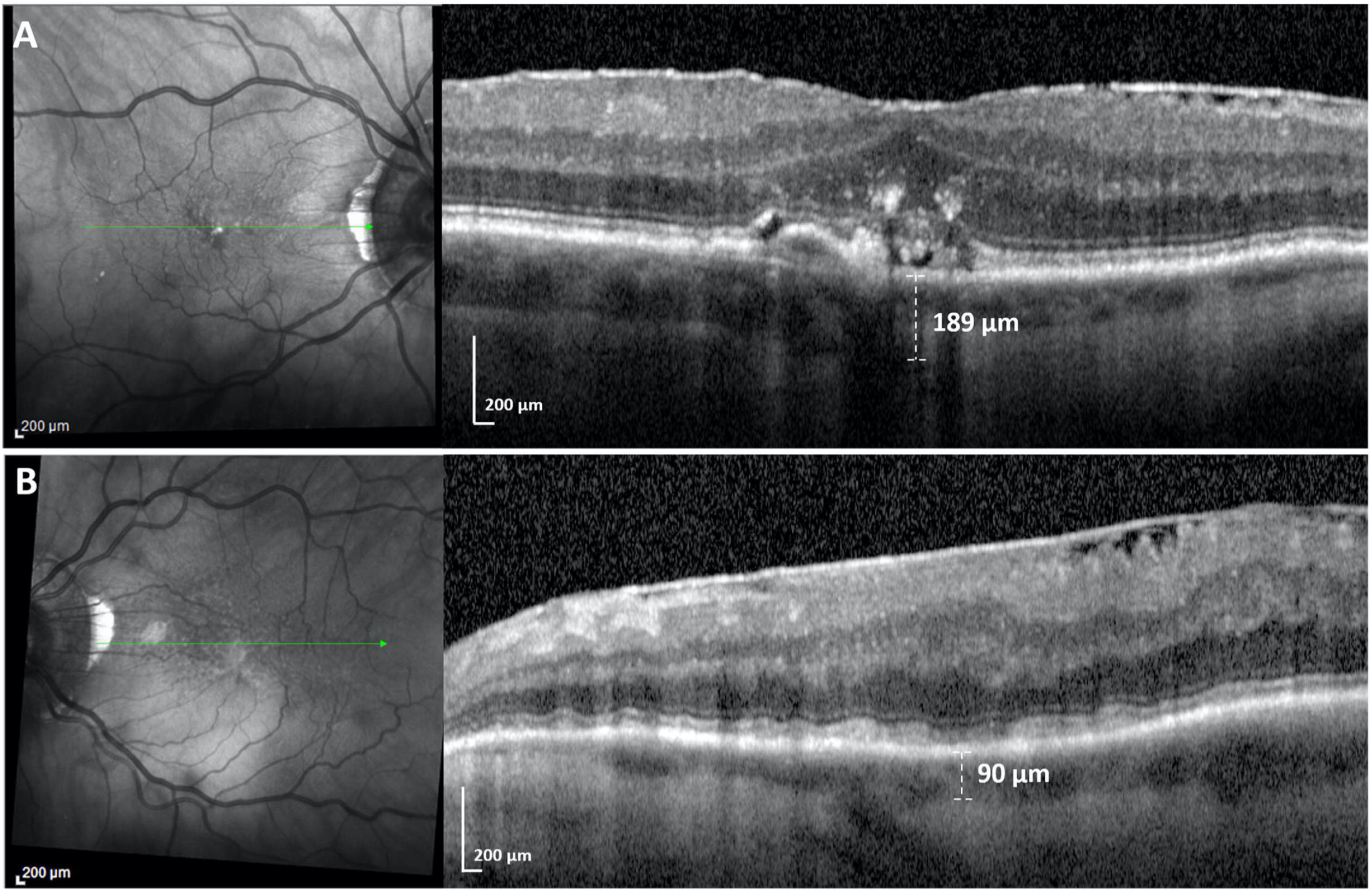
An experiment in Nature. Near Infra-red (NIR) + SD-OCT scan (both eyes) of a male in 81-85 year-old age range with AMD, unilateral left internal carotid artery (ICA) occlusion and carotid stroke. **A) Right eye**. Imaging was typical of AMD. NIR showed a few hyperreflective foci suggestive of drusen (green line, location of OCT scan). SD-OCT revealed confluent soft drusen and overlying retinal pigment epithelium (RPE) disruption. Subfoveal choroidal thickness was 189 microns. **B) Left eye**. NIR showed a well-defined group of multiple homogeneous hyporeflectant lesions, typical of SDD (green line, location of OCT scan). SD-OCT scan revealed multiple smooth confluent hyperreflective lesions (“ribbons” pattern) between the RPE and ellipsoid zone (EZ) and a subfoveal choroidal thickness of 90 microns. This is pure RMD (no soft drusen). Note that both structural features of RMD (SDD and choroidal thinning) are present purely and exclusively in the left eye, where the perfusion of the choroid by the ophthalmic artery is compromised due to internal carotid occlusion by plaque in the setting of CVA, supporting choroidal hypoperfusion as a mechanism for the presence of RMD in AMD, a perfect experiment in Nature in which occlusion of one ICA produced the exact RMD phenotype ipsilaterally, with no sign of it contralaterally. *\*Patient has bilateral epiretinal membranes, with foveal contour distortion in the left eye*.

**Figure 3.**
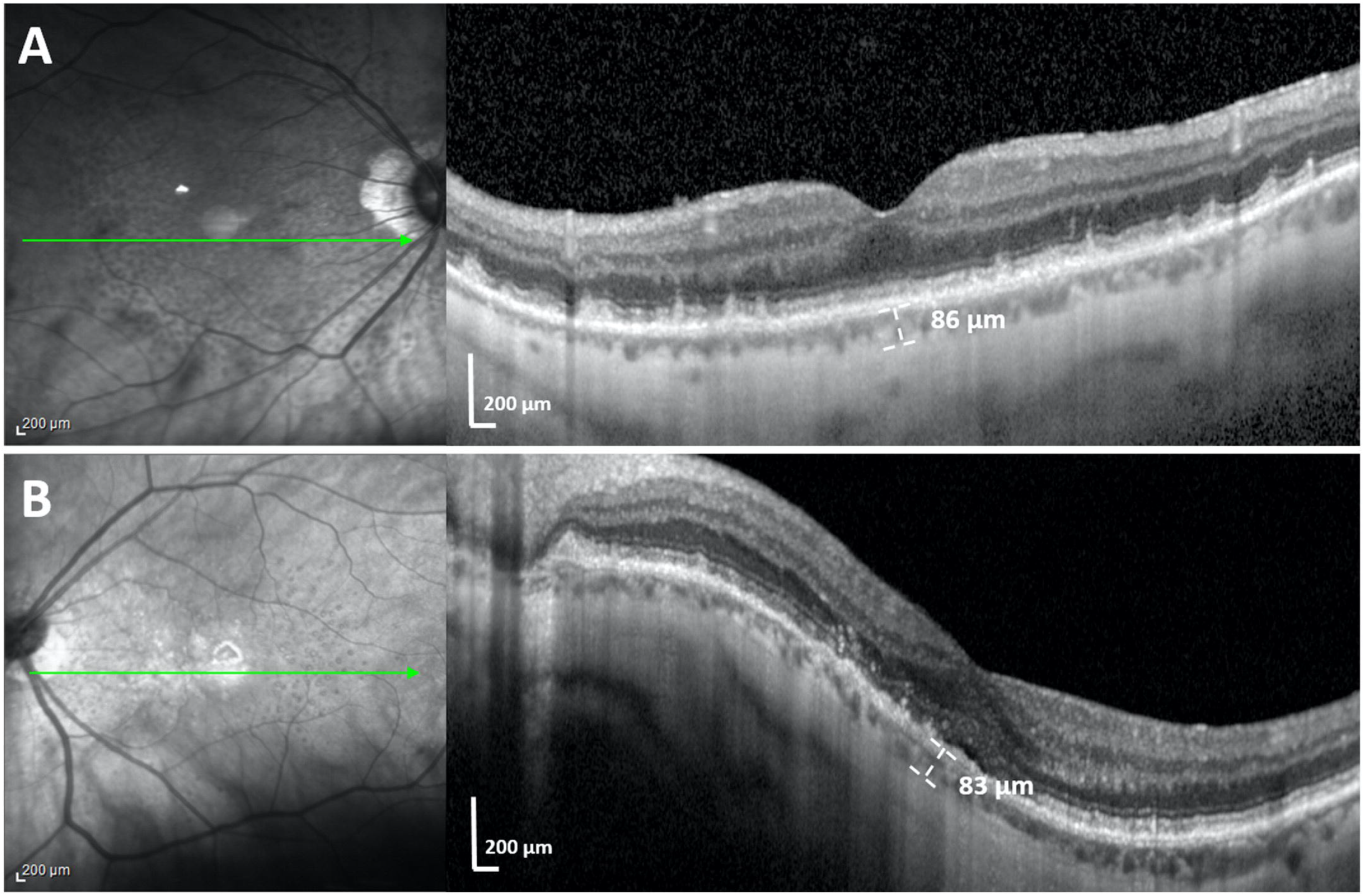
RMD and Cardiac Pump Deficiency. Near Infra-red (NIR) + SD-OCT scan (both eyes) of a male in 76-80 year-old age range with AMD, high myopia and history of Myocardial Infarction (MI). NIR scans showed well-defined groups of myriad homogeneous hyporeflectant lesions, typical of SDD (green lines, location of OCT scans). **A) Right eye**. SD-OCT revealed both structural components of RMD, a thin choroid of 86 microns and multiple hyperreflective subretinal lesions with a conical appearance (“dots” pattern) above the RPE, distorting (Stage 2), and penetrating (Stage 3), the ellipsoid zone (EZ). **B) Left eye**. SD-OCT revealed structural RMD, a thin choroid of 83 microns and multiple discrete hyperreflective lesions between the RPE and ellipsoid zone (EZ), again in a “dots” pattern. Few if any drusen are seen. Marked choroidal thinning OU in this subject is consequent to both high myopia and RMD. The myriad SDD OU, seldom seen in high myopia with choroidal thinning *per se*, are ascribed to the choriocapillaris insufficiency resulting from cardiac pump deficiency post MI.

**Figure 4.**
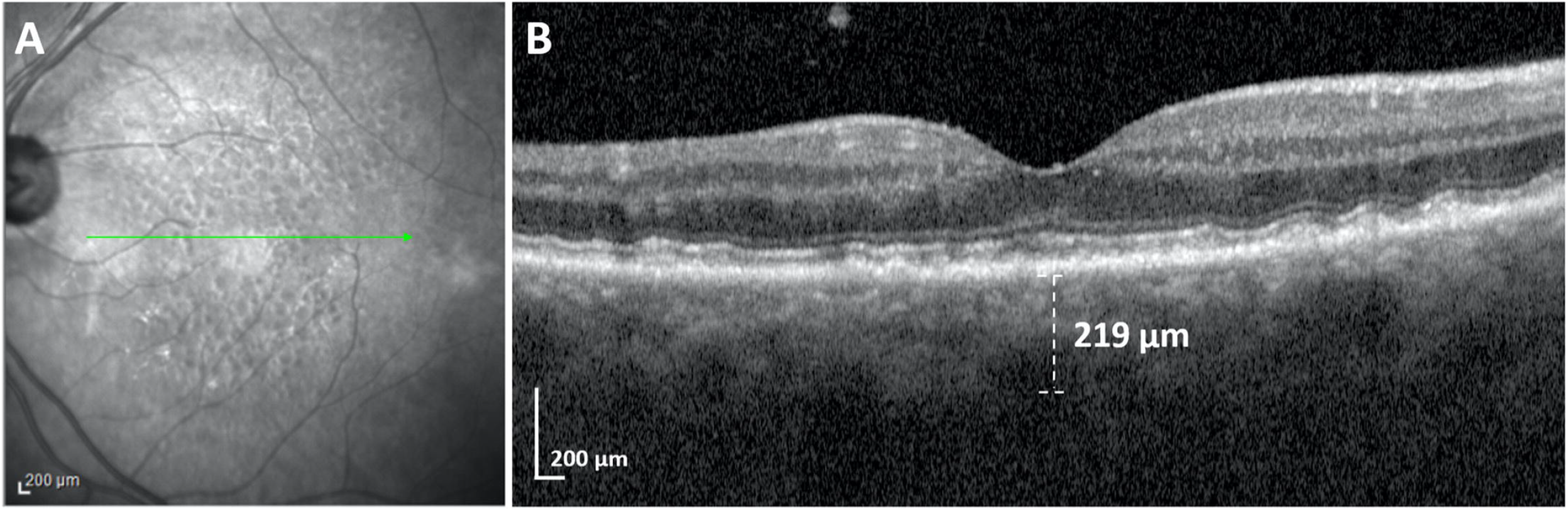
RMD and Cardiac Valve defect. Near Infrared (NIR) + SD-OCT scan (left eye) of a female in 81-85 year-old age range with RMD, Severe Aortic Stenosis, and no Atherosclerosis. **A)** NIR showed the typical SDD pattern of multiple hyporeflectant dots, some with hyper-reflectant centers suggesting distortion of the EZ (green line, location of OCT scan). **B)** SD-OCT scan revealed a pure RMD/SDD phenotype, multiple discrete hyperreflective lesions nasally (“dots”) with a conical appearance above the RPE distorting the EZ, and a smoother wave-like series of subretinal “ribbons” temporally, with no sign of drusen. The subject has 8 Diopters of hyperopia, which can produce ∼50 microns of choroidal thickening over emmetropic controls. The measured choroidal thickness of 219 microns corrected for refractive error is thus ∼169 microns, more typical for Reticular Macular Disease. *\*Right eye was not evaluable due to chronic Neovascular Glaucoma following an ischemic Central Vein Occlusion*.

Thus, herein, we prospectively evaluated AMD patients classified into RMD and nonRMD on the basis of presence or absence of SDD (with obligate underlying CC deficiency). Categories of High-Risk Vascular Disease (HRV) that increase risk of associated ophthalmic artery compromise were correlated with RMD status, serum cardiovascular risk markers, and demographic, ocular and medical data. RMD status was also correlated with HRV status and other covariates.

## Methods

This was a multicenter prospective study conducted at two tertiary vitreoretinal referral centers in New York City, USA: Vitreous Retina Macula Consultants of New York (LY, KBF), and Department of Ophthalmology, New York Eye and Ear Infirmary of Mount Sinai School of Medicine (MSSM) (RBR, RTS), from January 2019 to January 2021. The institutional review board of MSSM approved the study, which adhered to the tenets of the Declaration of Helsinki. ClinicalTrials.gov Identifier: NCT04087356.

### Inclusion Criteria

All patients older than 50 years, diagnosed with either or both of the RMD/SDD and soft drusen forms of AMD in either eye, who signed informed consent, and completed a study-related questionnaire, were included.

**Exclusion Criteria** were other retinal degenerations and vascular diseases, prior retinal surgery (except intravitreal injections), poor-quality imaging and/or inconclusive medical or macular diagnoses.

**Patient Evaluation** included best-corrected visual acuity, slit lamp biomicroscopy, and intraocular pressure (mmHg), serum for vascular disease risk factors, age, gender, race/ethnicity, relevant family history, smoking, medication, and medical/ocular history specifically covering stroke and these heart disease histories: MI, coronary artery bypass grafting (CABG), angina, arrhythmia, positive stress test, positive cardiac catheterization, stent, valve disease and CHF of any cause. This entire group was the CVD and Stroke group. The High-Risk Vascular Diseases (HRVs) had 3 sub-categories: two for heart disease, Pump Defect (MI, CABG, CHF) and Valve Defect; and one for carotid stroke. All others in the study were considered non-High Risk for OA compromise. The 55 total self-reported cases of heart disease or stroke were subsequently checked against medical records; 50 could be confirmed and 1 was corrected, leaving 51 cases in the CVD and Stroke group; the other 4 were removed from the group. Records of subjects with valve disease were further reviewed for specific valve(s) and valve defects (stenosis or insufficiency), as were those of stroke patients for source/type of stroke (carotid plaque or other), and laterality.

### Imaging

Volume SD-OCT scans (27 lines, automated retinal tracking, 16 scans averaged per line), and *en face* AF and NIR scans (both 30 degrees) centered on the fovea on the Heidelberg Spectralis HRA + OCT (Heidelberg engineering, Heidelberg, DE).

### Image Analysis

SDD grading on SD-OCT imaging for presence and most advanced stage followed a published protocol,^10^ independently and then to consensus by two retina specialists (GLG, OOM). Any differences were resolved by a senior grader (RTS). Stage 1 and 2 SDD, (**Figure 1D**). Stage 3 SDD, (**Figure 3A**). AF and NIR images (**Fig 1B, C**) were used to confirm the presence of SDD as fairly homogeneous hypoautofluorescent or hyporeflective lesions in compact distributions with well-defined borders.^16^ Soft drusen were identified on SD-OCT imaging as lesions of intermediate reflectivity on Bruch membrane (BrM) elevating the RPE (**Figure 1H**). Patients were assigned to 2 groups, SDD/RMD present (either eye, +/-drusen) or absent, nonRMD (soft drusen present, either eye). Choroidal thickness (CTh) was measured on a central SD-OCT scan (**Fig 1)**.

### Serum CVD Risk Markers Analysis

Blood samples for risk biomarkers of CVD^25^ (high-density lipoprotein cholesterol (HDL-c), low-density lipoprotein cholesterol (LDL-c), triglycerides (TG), total cholesterol (TC) and high sensitivity C-reactive protein (hsCRP)) were rapidly centrifuged at 1800 g for 10 minutes and refrigerated. Lipid levels were measured (Quest Diagnostics, Teterboro, NJ) by spectrophotometry, and plasma levels of hsCRP, by Immunoturbidimetric Assay (Orion Diagnostica, Finland). AMD risk genes were not analyzed in this study.^26^

### Statistical Analysis and Machine Learning

Experts in both fields (AB, AG) used ‘IBM SPSS Statistics version 27’, ‘Waikato Environment for Knowledge Analysis (WEKA) Version 3.8.5’, a data modeling tool, and Microsoft Excel 365. Univariate statistics were chi square for categorical variables, two-tailed t-test for continuous variables, with medians, ranges and/or interquartile ranges (IQRs) for non-normally distributed data. Multivariate regression determined each variable’s significance (p< 0.05) after controlling for all other covariates.

For bidirectional prediction of both HRV and RMD status, Logistic Model Trees (LMTs) were built from all covariates. ^27^ These exemplary models for representing the data (not uniquely determined like multivariate regression) included only variables of consequence, with less chance of overfitting and greater robustness to the data. Thus, variables of lower univariate significance, p-values > 0.1, were omitted *a priori*. Variables with minimal impact on performance were discarded iteratively. The resulting models were then tested by standard prediction statistics (accuracy, specificity and sensitivity with 95% confidence intervals (CIs)). LMT1 predicted RMD status. LMT2 predicted HRV status.

LMTs were also compared to multinomial logistic regression and an *ad hoc* logical model (LM) for HRV.

## Results

### Demographics and clinical characteristics

Of the 151 subjects with AMD, 75 had RMD present and 76 absent on imaging. 6 disagreements were resolved by consensus without arbitration. The associations of demographic, ocular and clinical characteristics of patients with or without RMD were not significantly different, except that Acetylsalicylic acid (ASA) use was significantly associated with RMD (53/75 RMD vs 30/76 nonRMD, p= 0.0001, OR 3.69, 95%CI 1.88-7.27), and mean CTh was less in RMD eyes than in nonRMD eyes (Mean 146±58 vs. 177±59 microns, p**=** 0.003; median 161.5, range 78-228 microns vs. median 177.5, range 90-286 microns).

### Serum CVD risk markers

Cholesterol panel and hsCRP mean values for RMD and non-RMD subjects were not significantly different, except mean HDL trended lower in RMD (mean 61±18, median 57, IQR 19) than non-RMD subjects (mean 69±22, p= 0.038, median 69, IQR 29). For risk of HRV, the discriminant HDL<62 mg/dL significantly correlated with HRV, and the standard lab cutoff HDL<40 mg/dL did not, HDL<62 was found in 24/29 HRV, 42/122 nonHRV, OR 12.40, 95% CI 5.125-30.014; p= 0.0002

### Vascular conditions

HRVs: 29 total of 151 subjects, 25/29 RMD, OR 9.0, CI 2.9 –27.46 (p= 0.000012). HRV subgroups, **Table 1**.

**Table 1.**
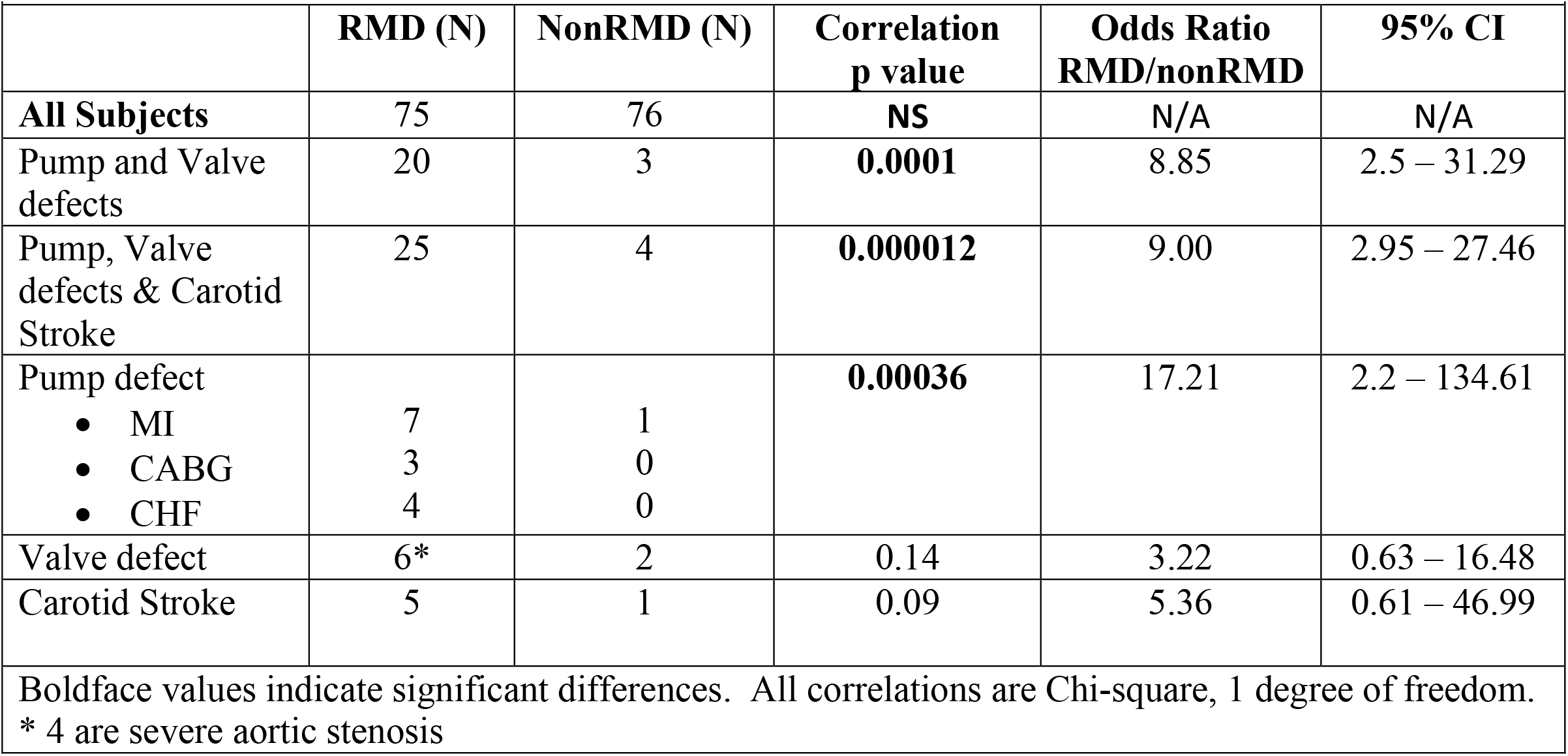
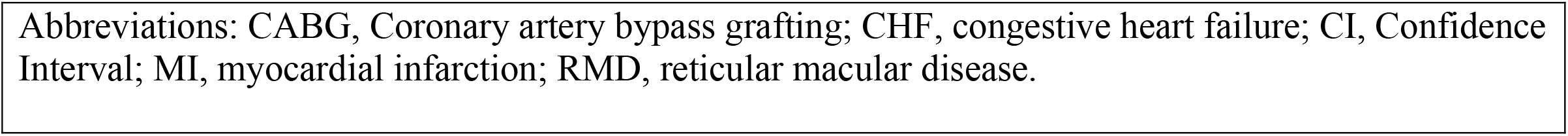
High Risk Vascular Diseases and Reticular Macular Disease: Correlations and Odds Ratios.

|  |  |  |  |  |  |
| --- | --- | --- | --- | --- | --- |
|  | **RMD (N)** | **NonRMD (N)** | **Correlation**<br>**p value** | **Odds Ratio**<br>**RMD/nonRMD** | **95% CI** |
| **All Subjects** | 75 | 76 | **NS** | N/A | N/A |
| Pump and Valve defects | 20 | 3 | **0.0001** | 8.85 | 2.5 – 31.29 |
| Pump, Valve defects & Carotid Stroke | 25 | 4 | **0.000012** | 9.00 | 2.95 – 27.46 |
| Pump defect |  |  | **0.00036** | 17.21 | 2.2 – 134.61 |
| • MI | 7 | 1 |  |  |  |
| • CABG | 3 | 0 |  |  |  |
| • CHF | 4 | 0 |  |  |  |
| Valve defect | 6* | 2 | 0.14 | 3.22 | 0.63 – 16.48 |
| Carotid Stroke | 5 | 1 | 0.09 | 5.36 | 0.61 – 46.99 |
| **Boldface values indicate significant differences. All correlations are Chi-square, 1 degree of freedom.**<br>**\* 4 are severe aortic stenosis** |  |  |  |  |  |

Abbreviations: CABG, Coronary artery bypass grafting; CHF, congestive heart failure; CI, Confidence Interval; MI, myocardial infarction; RMD, reticular macular disease.

Remainder of CVD and Stroke group: 24 total, 11 RMD, 13 nonRMD

Entire CVD and Stroke group, 53 total of 151 subjects, 36/75 RMD, 17/76 nonRMD, p= 0.0047, OR 3.20, 95%CI 1.58-6.48

### Multivariate models

Multivariate regression found only HRV status significant for RMD risk, (p= 0.000333) after controlling for all other parameters. For HRV risk, we found RMD status, ASA use and HDL<62 remained significant (p= 0.000098, 0.010 and 0.021, respectively).

For prediction, the bidirectional LMTs^27^ provided the most robust descriptions of the small dataset after serially removing variables of lesser significance. ASA use, a *treatment* for HRV, was rejected *a priori* in models for HRV risk, and similarly rejected for RMD risk, considering the evident strong association of HRV with RMD/SDD.

LMT1 for presence of RMD, given HRV status

1. RMD = -0.46 + [HRV=1] * 1.18 Accuracy 73.0 %, 95% CI (64.4% - 80.5%)

LMT2 for HRV status (not shown) was less effective than the following transparent logical model LM2.

2. IF (RMD= YES) AND (HDL<62) THEN (HRV=YES) LM2 achieved specificity 87.4%, sensitivity 77.4%, accuracy 84.9%; 95% CIs 79.0% -93.3%), 58.0%-90.4%, 77.5%-90.7%, respectively.

## Discussion

We have presented persuasive evidence that SDD/RMD in AMD eyes is a strong biomarker for co-existing HRVs: Cardiac (Pump Insufficiency, Valve Defects) and Carotid Stroke (with accompanying ICA plaque), whose mechanism is inadequate perfusion of the ophthalmic artery (OA) leading to CC insufficiency. This connection had eluded retinal research for decades in dozens of studies,^3-7^ even recent papers on SDD and choroidal abnormalities.^28,29^ although strong clues abound. SDD are associated with decreased longevity,^13^ with vascular death suspected^10,11^ Histopathology of 1777 donor eyes^30^ found SDD, alone among AMD lesions, were associated with cardiovascular death. OA flow on MRA was found decreased in AMD vs conrols.

The cholesterol-rich drusen and SDD^8^ superficially resemble atherosclerotic disease (ASD), which led to a search for common mechanisms. Drusen dynamics have now been correctly associated with local lipid pathways and associated genetics.^8^ The risk marker of low serum HDL for ASD actually runs contrary for AMD, with higher HDL increasing risk.^31^ This apparent paradox can now be explained: High HDL is only a risk for *drusen* in AMD;^31^ low HDL is a risk for ASD, which we now find is a risk for RMD, the other AMD pathway. HDL cuts both ways, depending on the pathway.

It is also clear that HRV and RMD, which share risk factors, do not share mechanisms, rather that HRV itself, with its own systemic mechanisms, is in turn the mechanism for much of RMD.

The logical model LM2 (**Eqn 2**), combining RMD status and a critical HDL level (HDL<62), predicts HRV with high accuracy, sensitivity and specificity, with immediate relevance to public health. The high specificity in particular suggests that a subject with RMD and lower HDL is high risk for HRV, and should be considered for **immediate, basic vascular work up**: EKG+/-stress test, cardiac and carotid echo for the 3 main classes of HRV. It is also of interest that the discriminant HDL<62 captured most of our HRV subjects, whereas the usual criterion for ASD risk of HDL<40 did not.

The clarity of the mechanism, and the many cases of pure RMD (without other AMD signs) suggest that RMD is not initially AMD, but is a major, distinct retinal pathology like diabetic retinopathy, driven by systemic vascular disease. Outer retinal damage then can flow, along with that of soft drusen, into AMD’s advanced forms, with similar presentations. But even the end stages of atrophic AMD, geographic atrophy, that ensue from drusen and SDD, have distinguishing autofluorescence characteristics.^32^ Examples of pure RMD are shown: with thin choroid (**Figure 1D)**, bilaterally (**Figure 3**) with MI (Pump Insufficiency); unilaterally (**Figure 2**), with stroke and ipsilateral ICA occlusion; with severe aortic stenosis (Valve Defect) and no ASD (**Figure 4**). Indeed, in another study, eyes with nearly pure SDD had 10-fold higher late AMD risk than similar early AMD eyes,^33^ suggesting that these worse prognosis eyes may be pure SDD phenotypes relentlessly driven by HRV.

Choroidal blood supply, from the posterior ciliary arteries (PCAs) supplied by the ophthalmic artery (OA), is a crucial component of the vascular pathway to RMD/SDD. It is an end-arterial system supplying CC lobules^9^ without adjacent anastomoses that are vulnerable to ischemia, while the retinal circulation derives protection by autoregulation.^34^ Further, although we found the expected significant choroidal thinning with RMD, choroidal thinning *per se* is not equivalent to CC insufficiency, but appears to be a sign of the generally poor choroidal perfusion that attends RMD in this context. Other vascular risk factors may also need re-evaluation. Smoking is a major risk for RMD,^13^ but the risk may actually be mediated through CAD and carotid stroke, rather than independent choroidal toxicity.

There is a wide breadth of applications for this new pathophysiological paradigm. The extraordinary preponderance of women (∼85%) among older AMD subjects with RMD^16^ can now be explained by the significantly earlier death of men^1^ than women from HRV;^1^ hence, as we previously pointed out,^10^ a strong linkage of HRV with RMD makes female preponderance of RMD in later years inevitable.

The study has several weaknesses. The sample size is relatively small, so the results, from a mostly Caucasian elderly population, invite replication in larger and diverse cohorts e.g., in Asians, with the prevalent polypoidal vasculopathy form of AMD.^35^ Another strong risk factor for SDD, discovered by our group^36^ but not considered here, is genetic: the Age-Related Maculopathy susceptibility (*ARMS2*) risk allele.^36^ *ARMS2* risk is likely acting independently of HRV to drive some cases of RMD, a testable hypothesis. Abnormal lipid trafficking in SDD formation and the association of HDL<62 with HRV are unexplained and merit investigation. Vascular histories were patient reported and were verified from medical records only in positive cases with stated heart disease or stroke. Severity of disease classifications or metrics such as cardiac ejection fraction, percent ICA occlusion or OA flow were not obtained. They would be very useful to interpreting these results, and should be included in future studies.

Strengths of this prospective study at two tertiary retina referral centers include rigorous patient selection and AMD phenotyping with high quality multimodal imaging for drusen and RMD. The extraordinarily high statistical significance of the associations found between HRV and RMD, contrasted with ambiguous findings from the world’s largest studies, suggests that the targeted subsets of AMD and vascular disease were the correct ones, and they were also convincingly associated by a perfectly fitting, single vascular mechanism. The bidirectional machine learning models also corroborated the connection: the logical model LM2 accurately predicts HRV from the presence of RMD and lower HDL, suggesting valuable, perhaps lifesaving, clinical applications.

## Conclusion

The presence of SDD/RMD lesions of AMD on retinal imaging and lower serum HDL cholesterol are an accurate predictor of certain high risk vascular diseases (HRVs). The mechanism is compromised perfusion of the outer retina. This opens the door for detection of HRV with standard, inexpensive testing. Further, potentially detectable HRVs compromising cardiac output include ASD-unrelated disorders like aortic stenosis and idiopathic CHF. There may be others.

Larger and detailed studies from both the ophthalmic and vascular perspectives are warranted to assess the full impact of these findings on detection of co-existing HRV. However, the accuracy of the prediction model for highly prevalent HRV, suggests that it will be significant. Larger studies in more diverse populations will also doubtless reveal major exceptions to and refinements of the paradigm, another important goal.

Predicting HRV from RMD (LM2) and RMD from HRV (LMT1) can help prevent mortality, morbidity and blindness from HRV and AMD. The care of HRVs could thus include retinal evaluation for RMD, with its high risk of AMD progression.^13-15^ Identification of RMD in eye clinics and large scale ophthalmic screening for SDD/RMD in underserved communities, with appropriate referral for systemic vascular work-up, could become a high impact public health initiative. In particular, detection of RMD in women should prompt evaluation for *undiagnosed* cardiac disease, a serious issue in women’s health.^37,38^

## Data Availability

N/A

## Acknowledgment

We gratefully acknowledge several helpful conversations with Christine Curcio.

## Notes

**Funding:** Regeneron Pharmaceuticals Investigator-Initiated Study, Research to Prevent Blindness Challenge Grant, Macula Foundation (KBF), Bayer-Global Ophthalmology Awards (GLG), International Council of Ophthalmology-Alcon Fellowship (OOM), New York Eye and Ear Infirmary Foundation (SA).

### Competing Interest Statement

Richard B. Rosen is a consultant to OptoVue, Boehringer-Ingelheim, Astellas, Genentech-Roche, NanoRetina, OD-OS, Regeneron, Bayer, Diopsys and Teva. He has personal financial interests in Opticology, Guardion and CellView. K. Bailey Freund is a consultant to Regeneron, Allergan, Zeiss, Bayer, Heidelberg Engineering and Novartis. He receives research funding from Genentech/Roche. R Theodore Smith is a consultant to Ora Technologies. The remaining authors have no relevant disclosures.

### Clinical Trial

NCT04087356

### Funding Statement

Regeneron Pharmaceuticals Investigator-Initiated Study, Research to Prevent Blindness Challenge Grant, Macula Foundation (KBF), Bayer-Global Ophthalmology Awards (GLG), International Council of Ophthalmology-Alcon Fellowship (OOM), New York Eye and Ear Infirmary Foundation (SA).

### Author Declarations

Regeneron Pharmaceuticals Investigator-Initiated Study, Research to Prevent Blindness Challenge Grant, Macula Foundation (KBF), Bayer-Global Ophthalmology Awards (GLG), International Council of Ophthalmology-Alcon Fellowship (OOM), New York Eye and Ear Infirmary Foundation (SA). This was a multicenter prospective study conducted at two tertiary vitreoretinal referral centers in New York City, USA: Vitreous Retina Macula Consultants of New York (LY, KBF), and Department of Ophthalmology, New York Eye and Ear Infirmary of Mount Sinai School of Medicine (MSSM) (RBR, RTS), from January 2019 to January 2021. The institutional review board of MSSM approved the study, which adhered to the tenets of the Declaration of Helsinki. ClinicalTrials.gov Identifier: NCT04087356.

